# Looking to and Processing of Audiovisual Speech and Associations with Language in Infant Siblings of Autistic and Non-autistic Children

**DOI:** 10.64898/2026.03.10.26347805

**Authors:** Kacie Dunham-Carr, Bahar Keçeli-Kaysılı, Jennifer E. Magnuson, Grace Pulliam, S. Madison Clark, Jacob I. Feldman, Pooja Santapuram, Kelsea McClurkin, Drina Agojci, Ava Schwartz, David J. Lewkowicz, Tiffany G. Woynaroski

## Abstract

Differences in looking to and processing of audiovisual speech have been theorized to contribute to heterogeneity in language ability in autistic children. Differential audiovisual speech processing has been indexed by event-related potentials (ERPs), specifically via amplitude suppression in response to audiovisual versus auditory-only speech, and linked with vocabulary in school-aged children. This study used an intact-group comparison and concurrent correlational design in infant siblings of autistic children (Sibs-Autism) and non-autistic children (Sibs-NA) to determine whether amplitude suppression is (a) present in infancy, (b) different in Sibs-Autism versus Sibs-NA, and (c) related to looking to audiovisual speech and language abilities. We collected EEG data from 54 infants aged 12-18 months (29 Sibs-Autism; 25 Sibs-NA) while they viewed videos of audiovisual and auditory-only speech, as well as eye tracking and language data. We found significant amplitude differences at the N2 ERP component in response to audiovisual versus auditory-only speech but no significant group differences in ERP amplitudes. Associations between looking to audiovisual speech, amplitude effects, and language were moderated by group, chronological age, and biological sex. Our findings suggest that differential audiovisual speech processing is present in 12–18-month-olds and may explain heterogeneity in looking to audiovisual speech and emerging language ability.

---

Autism is a neurodevelopmental condition with great heterogeneity in the presentation of features, the level of support needs, and long-term outcomes for affected individuals (Geschwind, 2009; Masi et al., 2017). Understanding the heterogeneity in language ability in autism is of particular interest and importance, as language ability by the age of five has been repeatedly associated with social, academic, and vocational outcomes for autistic^1^ individuals (e.g., Billstedt et al., 2007; Eisenberg, 1956; Gillberg & Steffenburg, 1987; Kobayashi et al., 1992). To further investigate heterogeneity in early development — including language development — pertaining to autism, researchers have begun to prospectively follow infant siblings of autistic children (Sibs-Autism; Rogers et al., 2009; Szatmari et al., 2016). Sibs-Autism are at elevated likelihood for autism diagnoses, as well as for difficulties in learning language, compared to infant siblings of non-autistic children (Sibs-NA; Marrus et al., 2018; Ozonoff et al., 2014, 2018; Zwaigenbaum et al., 2007). Thus, Sibs-Autism are a population rich in heterogeneity in both autism features and language ability with the potential to shed light on the mechanisms of typical and altered language development related to autism.

One theory of differential language development relevant to autism is the cascading effects framework, wherein early differences in the processing of sensory stimuli are purported to cascade onto the development of higher-order skills (Bradshaw et al., 2022; Cascio et al., 2016; Wallace et al., 2020). Prior studies have identified links between early sensory processing and a broad range of distal skills in Sibs-Autism and Sibs-NA (e.g., Damiano-Goodwin et al., 2018; Feldman et al., 2021, 2024; Gunderson et al., 2024; Wagner et al., 2018). Within this framework, it has specifically been proposed that the processing and perception of audiovisual speech may influence language development (e.g., Bahrick & Todd, 2012; Stevenson et al., 2018; Wallace et al., 2020). In accord with the cascading effects framework, this study represents a preliminary test of the hypothesis that children’s looking to audiovisual speech cues (i.e., to the mouth of a person speaking) impacts their processing of audiovisual speech, which may impact a child’s ability to understand and ultimately to use language (i.e., receptive and expressive language; see Figure 1).

**Figure 1.**
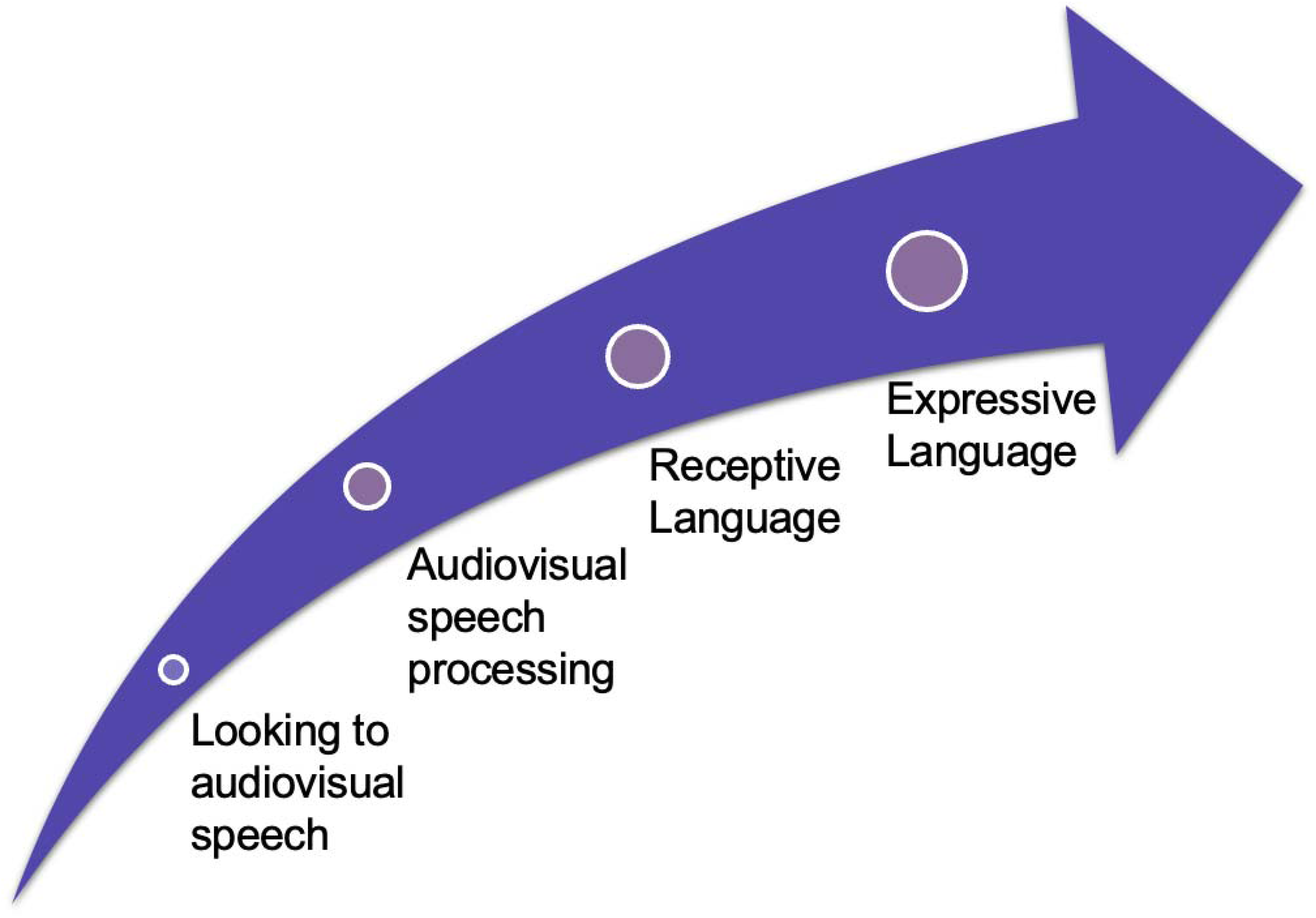
Overview of the Cascading Effects Framework as Applied to Looking to Audiovisual Speech. *Note.* An overview of the cascading effects framework of language development motivating this study, whereby differences in looking to audiovisual speech cues are theorized to explain variability in understanding and using language (i.e., receptive and expressive language) via audiovisual speech processing early in life.

## Looking to Audiovisual Speech and Links with Language Ability

Looking to audiovisual speech cues, specifically the moving mouth complementing the auditory speech stream as a source of multisensory redundant input, has been theoretically and empirically linked with language ability (see Bastianello et al., 2022 for a review). For example, eye tracking studies have identified an increase in looking to the mouth of a speaker in typically developing infants within developmental windows corresponding with qualitative changes in early language acquisition, such as the emergence of canonical babbling and the later spurt in word learning, around both 8-10 and 18 months of age (Hillairet de Boisferon et al., 2018; Lewkowicz & Hansen-Tift, 2012). Prior work has found that across Sibs-Autism and Sibs-NA, looking to the mouth of a speaker is indirectly associated with expressive language via multiple behavioral factors, including supported joint engagement with caregivers and the complexity of their prelinguistic vocalizations (Santapuram et al., 2022). Links with biological factors, including infants’ neural processing of audiovisual speech as described in the next section, may further explain variability in their early looking behavior and emerging language abilities.

## Differential Audiovisual Speech Processing as Measured by Electroencephalography (EEG)

Using EEG, differential audiovisual speech processing has been quantified via amplitude effects apparent in the event-related potential (ERP) in response to audiovisual speech versus auditory-only speech (van Wassenhove et al., 2005). Specifically, ERP amplitudes in response to audiovisual speech tend to be reduced, or suppressed, in non-autistic adults at the P2 (positive peak that occurs about 200 ms post-stimulus) component, which is purported to reflect early processing of sensory stimuli (van Wassenhove et al., 2005). P2 amplitude suppression in response to audiovisual versus auditory-only speech has been theorized to reflect multisensory integration of the phonetic features of auditory and visual speech (Baart et al., 2014; Kaganovich & Schumaker, 2014; Sato, 2024; Stekelenburg & Vroomen, 2007; van Wassenhove et al., 2005). P2 amplitudes in response to auditory-only and audiovisual speech are highly stable in autistic children (Dunham et al., 2020), and P2 amplitude suppression has also been observed in non-autistic and autistic children (Basirat et al., 2018; Dunham-Carr et al., 2023; Kaganovich & Schumaker, 2014; Knowland et al., 2014; Treille et al., 2017). Recent work shown that the degree of P2 amplitude suppression observed covaried with language ability in school-aged autistic children (i.e., with expressive vocabulary through receptive vocabulary in this population; Dunham-Carr et al., 2023).

## A Need to Extend Past Work to Understand Audiovisual Speech Processing in Infants

Less is known about ERP responses to audiovisual versus auditory-only speech in infants. Research focusing on auditory ERPs in response to speech in infants from the general population as young as 6 months and as old as 20 months has, however, identified responses at P1 (a positive peak occurring about 150 ms post-stimulus) and N2 (a negative deflection occurring between approximately 250-550 ms post-stimulus) components, with P1 likely reflecting detection and processing of acoustic stimuli and N2 likely reflecting phonetic processing of speech (e.g., Rivera-Gaxiola et al., 2007, 2012; Zhang et al., 2011). One investigation has reported that 6-9-month-old infants demonstrate a positive peak occurring about 150 ms post-stimulus (140-240 ms) in response to not only auditory-only speech but also congruent and incongruent audiovisual speech syllables. This study provided preliminary support for the possibility that amplitudes in response to audiovisual speech may be reduced, on average, relative to amplitudes in response to auditory-only speech at this component (see supplemental materials from Kushnerenko et al., 2013). Thus, it is possible that ERP amplitude effects to audiovisual versus auditory-only speech are present during the first and second years of life at varying ERP components, though research has been limited to date.

## A Need to Consider Participant Characteristics

Notably, past studies providing empirical support for the cascading effects of looking to and processing of audiovisual speech onto language acquisition pointed towards participant characteristics that may influence effects of interest. For example, past studies evaluating links between early sensory processing and behavior in Sibs-Autism and Sibs-NA have found associations moderated by sibling group (e.g., Damiano-Goodwin et al.; Feldman et al., 2024; Wagner et al., 2018) and chronological age (e.g., Feldman et al., 2021; Gunderson et al., 2024). Additionally, patterns of looking to audiovisual speech and links with language have been found to differ by biological sex in autistic children and infants at an elevated likelihood for autism (e.g., Chawarska et al., 2016; Harrop et al., 2020). Based on these findings from the extant literature, it is likely that links between audiovisual speech processing and language are also influenced by participant characteristics.

## The Current Study

This study sought to extend prior work on links between looking to audiovisual speech and language in Sibs-Autism and Sibs-NA, as well as past studies on associations between differential audiovisual speech processing and language ability in autistic and non-autistic children, by evaluating whether ERP amplitude effects in response to audiovisual versus auditory-only speech are present by 12-18 months of age, differ between Sibs-Autism and Sibs-NA, and covary with looking to audiovisual speech and language. We also considered the potential role of participant characteristics in testing associations in the current investigation. To our knowledge, this is the first study to investigate a neural proxy of differential audiovisual speech processing in Sibs-Autism. Our research questions were:

1. Are ERP amplitude effects in response to audiovisual versus auditory-only speech present across 12–18-month-old Sibs-NA and Sibs-Autism infants?
2. Are ERP amplitude effects for audiovisual versus auditory-only speech reduced, on average, in Sibs-Autism compared to Sibs-NA?
3. Are ERP amplitude effects associated with concurrent looking to audiovisual speech as measured by eye tracking?
4. Do eye tracking metrics of looking to audiovisual speech and ERP amplitude effects covary with concurrent receptive and expressive language?
5. Do participant characteristics influence relations between looking to audiovisual speech, amplitude effects, and emerging receptive and expressive language?

We hypothesized that differences in ERP amplitudes in response to audiovisual speech versus auditory-only speech would be present by 12-18 months of age, but that Sibs-Autism would show differences in ERP amplitudes that were smaller in magnitude compared to Sibs-NA. We also hypothesized that ERP amplitude effects would be related to looking to audiovisual speech, and that looking to audiovisual speech and ERP amplitude effects would covary with concurrent receptive and expressive language. Finally, we hypothesized that some of the associations of interest would likely vary according to participant characteristics, including chronological age, likelihood group, and/or biological sex.

## Methods

### Participants

The study sample comprised a total of 54 12-18-month-old infants, 29 Sibs-Autism and 25 Sibs-NA. The two groups were matched on biological sex and chronological age. The infants in the current sample were drawn from a larger NIDCD-funded study of early sensory function and broader developmental outcomes at Vanderbilt University and, thus, our sample partially overlaps with samples in prior reports (e.g., Feldman et al., 2021, 2024; Markfeld et al., 2023). Eligibility criteria for both groups were (a) full-term birth, (b) no concomitant genetic or neurological disorders, (c) monolingual English-speaking household, and (d) at least one older sibling. Infants in the Sibs-Autism group were required to have at least one older sibling with an autism diagnosis by a licensed clinician according to DSM-5 criteria (American Psychiatric Association, 2013); diagnosis was required to be confirmed by either a member of the research team reviewing the prior record of the older sibling or a research-reliable administration of the Autism Diagnostic Observation Schedule, 2^nd^ edition (ADOS -2; Lord et al., 2012) and clinical interview conducted in our laboratory^2^. Infants in the Sibs-NA group were required to have (a) only non-autistic older siblings, as confirmed by screening below the threshold for autism concern (i.e., < 15) on the Social Communication Questionnaire (Rutter et al., 2003)^3^, (b) no first-degree relatives diagnosed with autism, and (c) no prior history or present indicators of developmental delays or disorders per caregiver report. The Vanderbilt University Institutional Review Board approved recruitment and study procedures. Caregivers provided written informed consent, and families were compensated for their participation. See Table 1 for a description of participant characteristics.

**Table 1.**
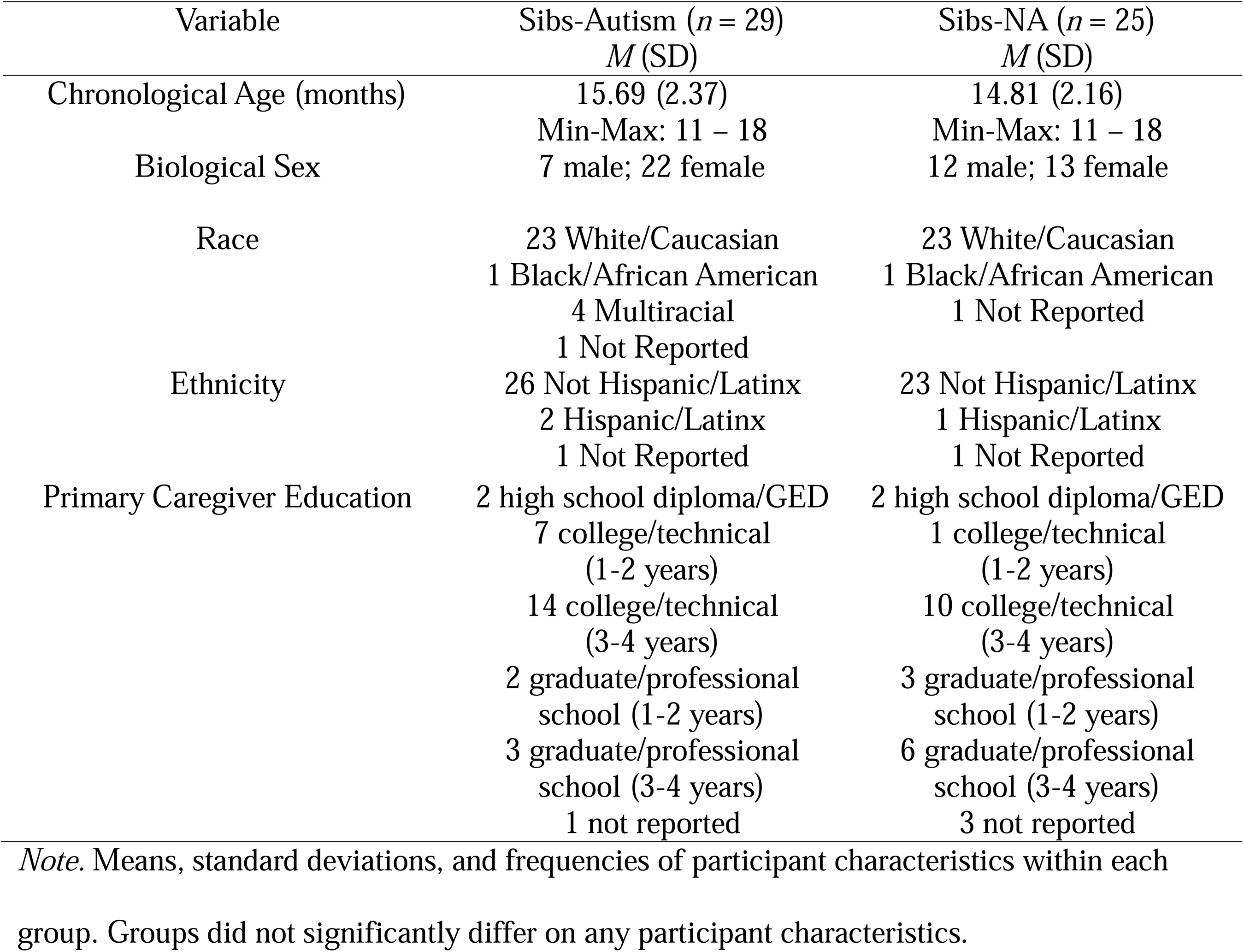
Description of Participant Characteristics According to Group.

### Electroencephalographic Measurement of Differential Audiovisual Speech Processing

#### Stimuli

The stimuli utilized in the ERP task consisted of the syllable “ba” as naturally spoken by a female speaker with a neutral facial expression against a white background. In the audiovisual condition, the corresponding auditory and visual stimuli were presented in synchrony, with visual speech cues preceding auditory speech onset as naturally produced by the speaker. In the auditory-only condition, the auditory stimuli were presented with a still image of the speaker’s face. These stimuli were adapted from prior investigations in our laboratory (Dunham et al., 2020; Dunham-Carr et al., 2023; Feldman et al., 2020). Images of baby toys (e.g., a train, a rattle) were presented periodically between trials (i.e., after every fourth trial, there was a 50% chance of a toy image appearing) to maintain participant attention to the monitor. All stimuli were presented via E-Prime.

#### EEG Data Collection Procedures

EEG data were collected using Net Station EEG Software and a 128-channel Geodesic sensor net (Net Amps 400 amplifier, Hydrocel GSN 128 EEG cap, EGI Systems Inc.). The raw signal was sampled at 1000 Hz and referenced to vertex (Cz), and impedances were kept at or below 40 kΩs. Data were collected while 50 trials of each stimulus type (i.e., audiovisual and auditory-only) were presented in random order. Trials were separated by an interstimulus interval that was randomly jittered between 400 ms and 850 ms. A member of the research team monitored participant attention throughout data recording and paused the task if the infant’s gaze deviated from the monitor.

#### EEG Data Processing

EEG data were bandpass filtered from 0.5 – 50 Hz, and bad channels and artifacts were manually removed in EEGLAB (Delorme & Makeig, 2004). Bad channels were interpolated, and data were re-referenced to average. Trials were baseline-corrected from 200 ms to 0 ms pre-stimulus onset. An average of 68 trials was retained after data cleaning, with an average of 33 auditory-only and 35 audiovisual trials per participant; the average number of trials retained did not significantly differ by group or condition (*p* values > .05) and was similar to prior work investigating ERPs in response to audiovisual speech in infants (Kushnerenko et al., 2013).

A temporal principal component analysis (PCA) was conducted across groups and conditions to confirm time windows of ERP components of interest for analyses, which were then applied to derive mean amplitudes from confirmed components of interest for each participant (Scharf et al., 2022). The temporal PCA utilized a six-factor solution and Promax rotation. The six-factor solution explained 82.58% of the variance, and the two factors with the highest loadings centered around 390 ms post-stimulus and 137 ms post-stimulus, respectively. These PCA results are comparable to those reported in similar studies investigating ERP responses to speech syllables in infants (Rivera-Gaxiola et al., 2005, 2007) and supported analyzing a P1 ERP component within a window 100-200 ms post-stimulus, as well as an N2 ERP component within a window 350-500 ms post-stimulus. Peak amplitudes from both components as measured at Cz were derived for use in analyses.

### Eye Tracking Measurement of Looking to Audiovisual Speech

#### Stimulus

The stimulus utilized in the remote eye tracking task used to measure looking to audiovisual speech was a 50 second video clip that has been used in several prior investigations of looking to audiovisual speech cues in infants and toddlers (e.g., Lewkowicz & Hansen-Tift, 2012; Pons et al., 2019; Santapuram et al., 2022). In the video clip, an adult female speaker produced a short monologue in English using infant-directed speech (i.e., with high pitch excursions, exaggerated prosody, and slow articulation, while smiling). The video was presented on a 24-inch computer monitor, with sound presented at 75 dB through an M-AUDIO BX8 D2 speaker placed in front of the participant just below the computer monitor. A Sensorimotorics Instrument (SMI) REDn Scientific Eye Tracking System (SMI, Teltow, Germany) was used to control stimulus presentation and to track eye gaze via pupil-centered corneal reflection.

#### Eye Tracking Data Collection and Processing Procedures

Participants were seated in a highchair or on their caregiver’s lap (with the caregiver’s eyes closed or covered via a pair of provided sunglasses throughout the procedure) 60-80 cm in front of the computer monitor, eye tracker, and speaker. Eye gaze was calibrated using a five-point calibration procedure. First, a brief video of a dancing Elmo cartoon was displayed to encourage attention to the monitor. Then, a looming star that moved from the center to each corner of the monitor was presented. The video clip was presented following eye gaze calibration. SMI’s BeGaze software was utilized to quantify gaze duration to a priori specified areas of interest (AOIs; the mouth, eyes, and face of the speaker, as detailed in prior work). A minimum of 5 seconds of looking to the face was required for data to be analyzed. Areas of interest were imposed on the speaker’s eyes, mouth, and face as described in Santapuram et al., 2022. The proportion of total looking time displayed to the mouth out of the total time spent fixating on any part of the face were derived for use in analyses. Analyses focused on the proportion of total looking time displayed to the eyes are included in Supplemental Materials.

### Measurement of Receptive and Expressive Language

#### Mullen Scales of Early Learning (Mullen)

The Mullen (Mullen, 1995) is a standardized developmental assessment for children birth-68 months of age. From this measure, we collected the Visual Reception, Fine Motor, Receptive Language, and Expressive Language scales. The age equivalency scores from the Receptive Language and Expressive Language scales were derived for analyses.

#### MacArthur-Bates Communicative Development Inventory (MCDI)

The MCDI (Fenson et al., 2007) is a caregiver report measuring early vocabulary acquisition. We calculated raw scores indexing the number of words a child “understands” (i.e., receptive vocabulary) and the number of words a child “understands and says” (i.e., expressive vocabulary) from the MCDI: Words and Gestures checklist for analyses.

#### Vineland Adaptive Behavior Scales, Second Edition (VABS-2)

The VABS-2 (Sparrow et al., 2005) is a caregiver report measuring a broad range of adaptive behavior from birth through age 90. The age equivalency scores from the receptive and expressive communication scales of this measure were derived for analyses.

## Data Analytic Approach

Discrete missing data (0-6.8% across all variables relevant to analyses) were imputed using the *missForest* package (Stekhoven & Bühlmann, 2012) in R (R Core Team, 2024). Data were missing due to technical difficulties, missed questions on parent reports, or the addition of study measures during data collection relevant to the larger project; thus, data were considered missing at random (Enders, 2010). All variables to be used in analyses were evaluated for normality, specifically for skewness >|1.0| and kurtosis >|3.0|. The Mullen Receptive Language age equivalent score and the MCDI Understands raw score were transformed via square root transformation, and the MCDI Says raw score was transformed via logarithmic transformation to correct for positive skew prior to aggregation and analyses.

### Creation of Language Aggregates

Pearson correlations were utilized to evaluate the intercorrelation between language indices purported to tap the same construct to ensure there was adequate empirical support for creating aggregated language scores. All indices purported to reflect receptive and expressive language, respectively, were sufficiently intercorrelated (*r* values for correlations between all component variables intended to tap the same construct ≥ .4; see Supplement) to warrant aggregation following *z*-score transformation (Rushton et al., 1983). Thus, a receptive language aggregate was calculated from the (square root-transformed) Receptive Language age equivalency score from the Mullen, the (square root-transformed) Understands raw score from the MCDI, and the Receptive Communication age equivalency score from the VABS-2. An expressive language aggregate was calculated from the Expressive Language age equivalency score from the Mullen, the (log-transformed) Understands and Says raw score from the MCDI, and the Expressive Communication age equivalency score from the VABS-2.

### Statistical Analyses

To answer our first and second research questions regarding ERP amplitude effects in response to audiovisual versus auditory-only speech across and between groups, we conducted a 2 (Group) x 2 (Condition) ANOVA to compare P1 and N2 amplitudes in response to audiovisual versus auditory-only speech. Partial eta squared values of .01, .06, and .14 indicated small, medium, and large effect sizes of these analyses. For our third, fourth, and fifth research questions regarding associations with looking to audiovisual speech and language, we first calculated P1 and N2 amplitude effects as the difference between the component amplitude in the auditory-only condition and the audiovisual condition (i.e., auditory-only P1 amplitude−audiovisual P1 amplitude and auditory-only N2 amplitude–audiovisual N2 amplitude). We then conducted a series of correlation and regression analyses to evaluate associations between P1 and N2 amplitude difference scores, looking to audiovisual speech cues as measured by eye tracking, and receptive and expressive language aggregate scores.

Moderation analyses were carried out using PROCESS in R (Hayes, 2022; R Core Team, 2024). Sibling group, chronological age, and biological sex were systematically evaluated as moderators of associations and retained in regression models when they were statistically significant. Interaction effects from these analyses were probed at *p* ≤ .1. This slightly lower Type I error threshold is often utilized for flagging significant interaction effects by default in statistics programs (e.g., R Core Team, 2024) and is commonly utilized in testing moderated effects in the autism literature (e.g., Feldman et al., 2021, 2024; Sandbank et al., 2020) to decrease the risk of making Type II errors, as interaction effects are difficult to detect in small samples (Aiken & West, 1991; Fairchild & MacKinnon, 2009). For continuous putative moderators (i.e., chronological age), we utilized Johnson-Neyman tests to probe and interpret significant moderation models. Cook’s *D* was utilized to monitor for the presence of outliers (defined as a Cook’s *D* value ≥ 1); no participants were identified as an outlier in any analyses. Detailed results of all significant regression models are provided in Supplemental Materials.

## Results

### Research Questions 1 and 2: ERP Responses to Audiovisual versus Auditory-only Speech Across and Between Groups

At P1, the 2 (Group) x 2 (Condition) ANOVA yielded no significant effects of Condition, *F*(1,52) = 1.09, *p* = .300, 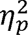 = .010; Group, *F*(1,52) = .661, *p* = .418, 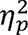 = .006; or Group x Condition, *F*(1,52) = .408, *p* = .524, 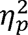 = .004. These effects were negligible to small in magnitude. At N2, however, the 2 (Group) x 2 (Condition) ANOVA yielded a significant effect of Condition, *F*(1,52) = 4.26, *p* = .042, 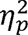 = .040, such that N2 amplitude was significantly more negative in response to audiovisual versus auditory-only speech, on average, across groups. This statistically significant effect was small in magnitude. There was not a significant between-group difference in N2 amplitude, on average, *F*(1,52) = .431, *p* = .513, 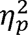 = .009, or a significant Group x Condition interaction, *F*(1,52) = .158, *p* = .692, 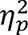 = .002. These non-significant effects were negligible to small in magnitude. See Figure 2 for a depiction of grand average ERPs across and within groups.

**Figure 2.**
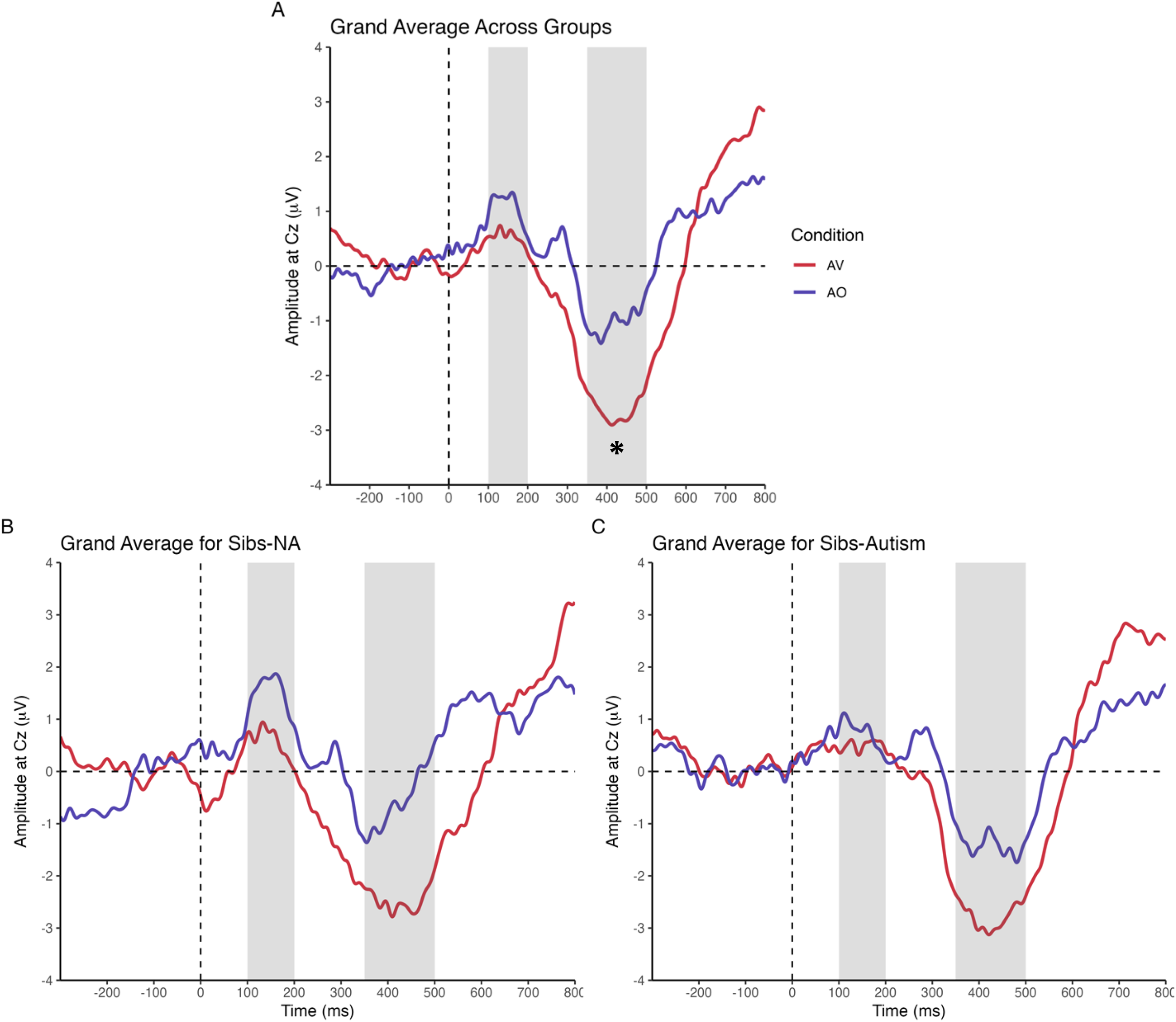
Grand Average Waveforms Across and Within Groups. *Note.* Grand average ERP waveforms for (A) all participants, (B) Sibs-NA infants, and (C) Sibs-Autism infants. Red lines illustrate the audiovisual (AV) response; blue lines illustrate the auditory-only (AO) response. Gray bars depict time windows of interest for P1 (100-200 ms post-stimulus) and N2 (350-500 ms post-stimulus), respectively. *\*p* value for effect < .05.

### Research Question 3: Associations Between Amplitude Effects and Looking to Audiovisual Speech

Associations between looking to the mouth and P1 amplitude effects varied according to group (*p* values for looking to mouth x group product term in moderation model = .017). Post hoc probes of these statistically significant group moderations revealed a moderate, positive association between looking to the mouth and P1 amplitude effects in the Sibs-Autism group (*r* = .328) and a moderate, negative correlation in the Sibs-NA group (*r* = -.333). Associations between looking to the mouth and P1 amplitude effects were not moderated by chronological age or biological sex (*p* values for interaction terms in regression models testing moderated effects >.1).

Associations between looking to the mouth and N2 amplitude effects (*p* value for looking to mouth x group product term in the moderation model = .053) also varied according to group. Post hoc probes of these group moderations revealed a moderate, negative correlation between looking to the mouth and N2 amplitude differences in the Sibs-NA group (*r* = -.427) and a negligible correlation in the Sibs-Autism group (*r* = .070). Associations between looking to the mouth and N2 amplitude effects were not moderated by chronological age or biological sex (*p* values for interaction terms in models testing moderated effects > .1). See Table 2 for a summary of regression models testing associations between looking to audiovisual speech and ERP amplitude effects and Figure 3 for a depiction of relations between looking to audiovisual speech and ERP amplitude effects according to group.

**Table 2.**
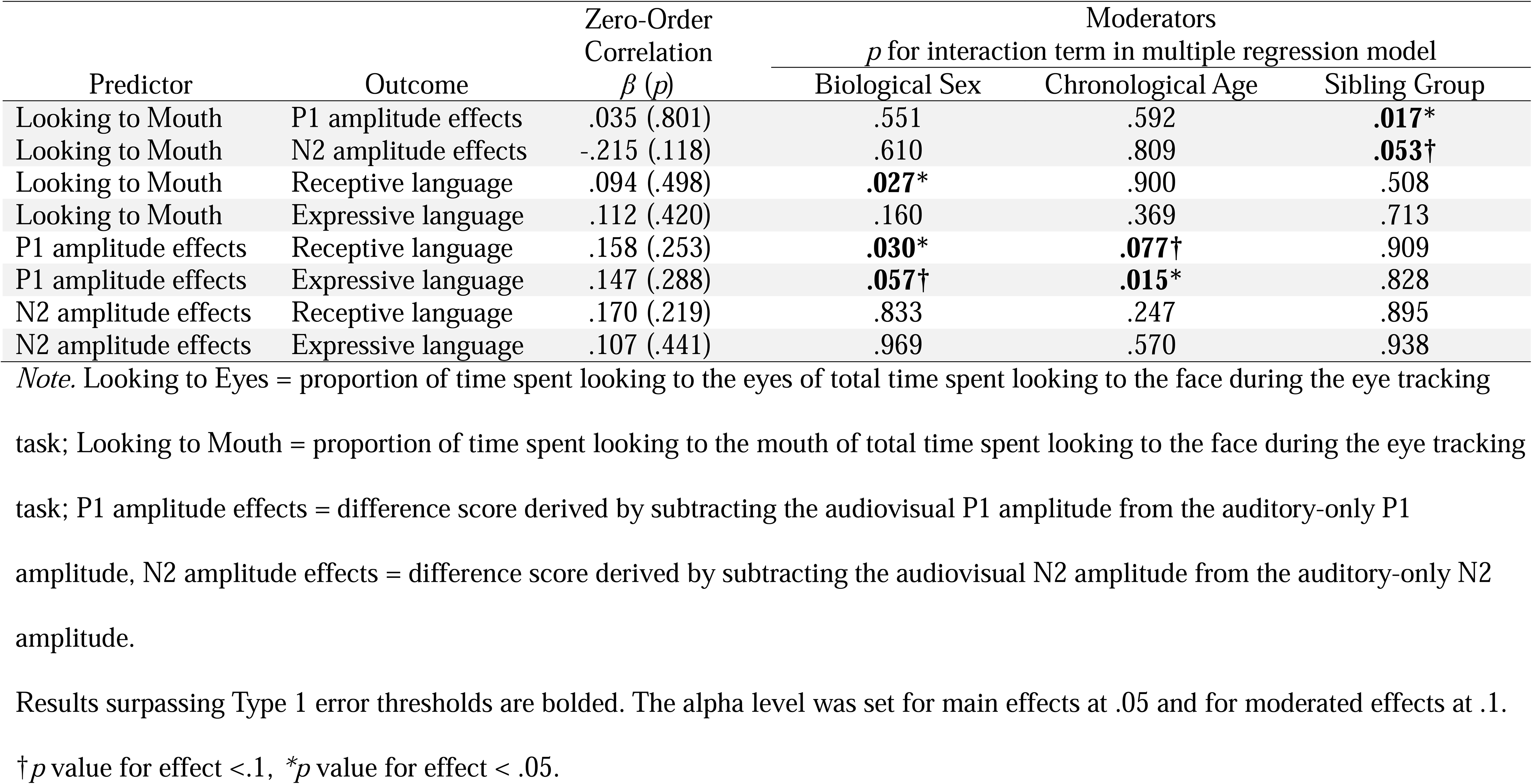
Summary of Key Correlation and Regression Results.

**Figure 3.**
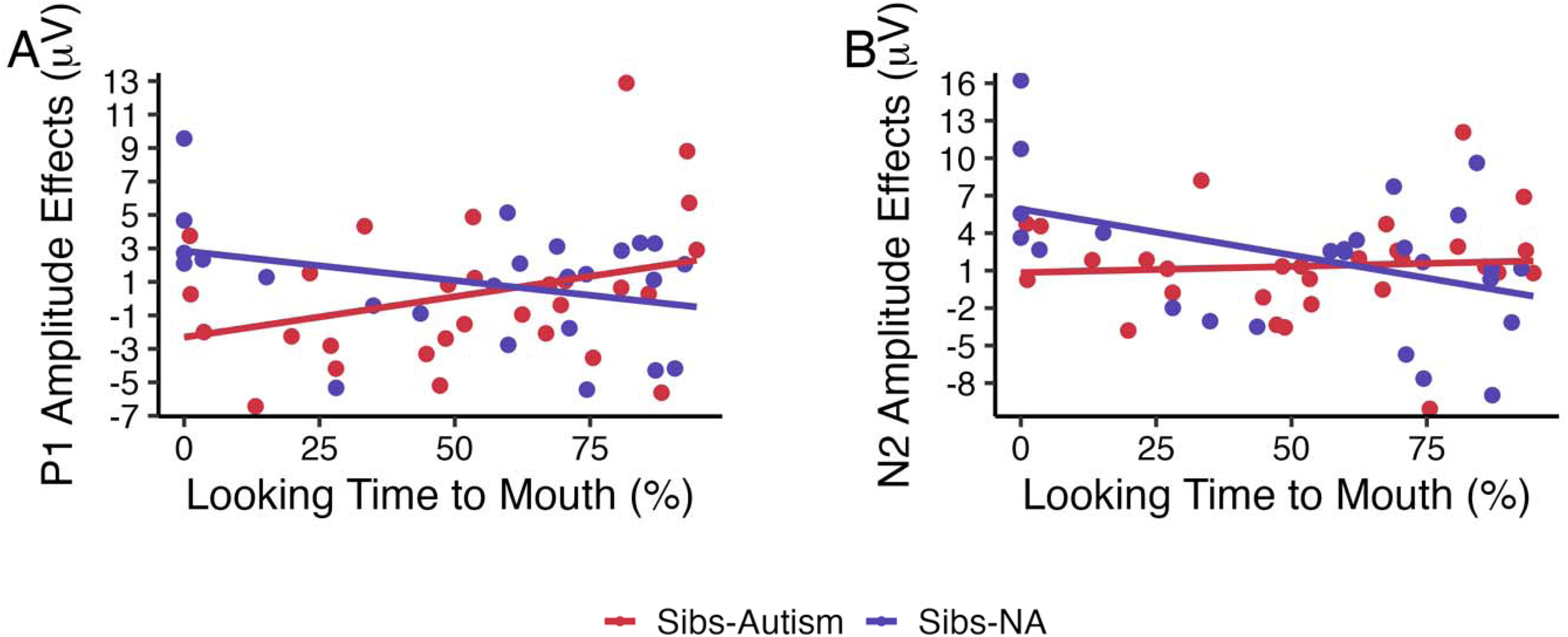
Relations between Looking to Audiovisual Speech and ERP Amplitude Effects According to Group. *Note.* Scatterplots depicting associations between looking to audiovisual speech and P1 and N2 amplitude effects, operationalized by the difference between the auditory-only and audiovisual amplitude. Associations were moderated by group, such that a greater proportion of time spent looking to the mouth corresponded with greater amplitude effects in Sibs-Autism but lesser effects in Sibs-NA. Red = Sibs-Autism group; blue = Sibs-NA group.

### Research Question 4: Associations Between Looking to Audiovisual Speech and Language

Associations between looking to the mouth and expressive language varied according to biological sex (*p* value for looking to mouth x biological sex product term in the moderation model = .027). Post hoc probes of this significant moderation by sex revealed a moderate, positive association between looking to the mouth and expressive language in males (*r* = .458) and a small, negative association between looking to the mouth and expressive language in females (*r* = -.150). This moderation was not statistically significant for the association between looking to the mouth and receptive language (*p* value for looking to mouth x biological sex product term in the moderation model = .160). Associations between looking to the mouth and receptive and expressive language were not moderated by group or chronological age (*p* values for interaction terms in models testing moderated effects > .1). See Table 2 for a summary of model results and Figure 4 for a depiction of regression results for relations between looking to audiovisual speech and receptive and expressive language according to biological sex.

**Figure 4.**
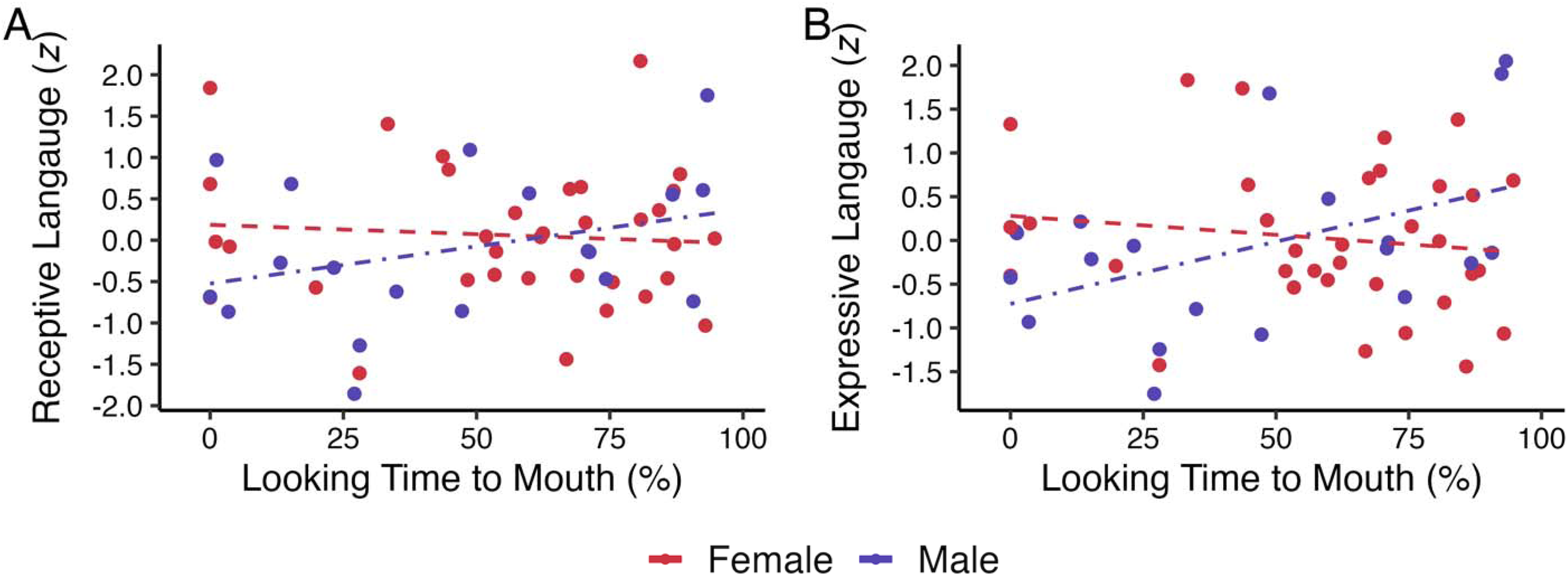
Relations between Looking to Audiovisual Speech and Receptive and Expressive Language According to Biological Sex. *Note.* Scatterplots depicting associations between looking to audiovisual speech and receptive and expressive language, as indexed by aggregate *z* scores. A greater proportion of time spent looking to the mouth was associated with higher receptive and expressive language scores in males and lower scores in females. Red = females, blue = males.

### Research Question 4: Associations Between ERP Amplitude Effects and Language

Chronological age was a moderator in the association between P1 amplitude effects and receptive language (*p* value for P1 amplitude effects x chronological age interaction term in the model testing moderation = .077). A Johnson-Neyman test was conducted to derive the precise cut-point along the moderator; relations between P1 amplitude effects and receptive language were significant beyond 14.03 months of age, such that greater P1 amplitude effects covaried with greater receptive language beyond this chronological age. Associations between P1 amplitude effects and receptive language also varied according to biological sex (*p* value for P1 amplitude effects x biological sex interaction term in the moderation model = .030). Post hoc probes identified a moderate, positive association between P1 amplitude effects and receptive language in males (*r* = .529) and a negligible association in females (*r* = -.026).

Associations between P1 amplitude effects and expressive language were significantly moderated by chronological age (*p* value for P1 amplitude effects x chronological age interaction term in the model testing moderation = .015). Johnson-Neyman test results indicated that the relation between P1 amplitude effects and expressive language was significant and positive in infants beyond 14.19 months of age. Biological sex also moderated associations between P1 amplitude effects and expressive language (*p* value for P1 amplitude effects x biological sex in the model testing moderation = .057), with a moderate, positive association between P1 amplitude effects and expressive language in males (*r* = .450) and a negligible association in females (*r* = -.009). Associations between P1 amplitude effects and receptive and expressive language were not moderated by group (*p* values for product terms in models tested moderated effects > .1). See Table 2 for a summary of results for tests of relations between P1 amplitude effects and language according to chronological age and biological sex and Figure 5 for a depiction of these associations.

**Figure 5.**
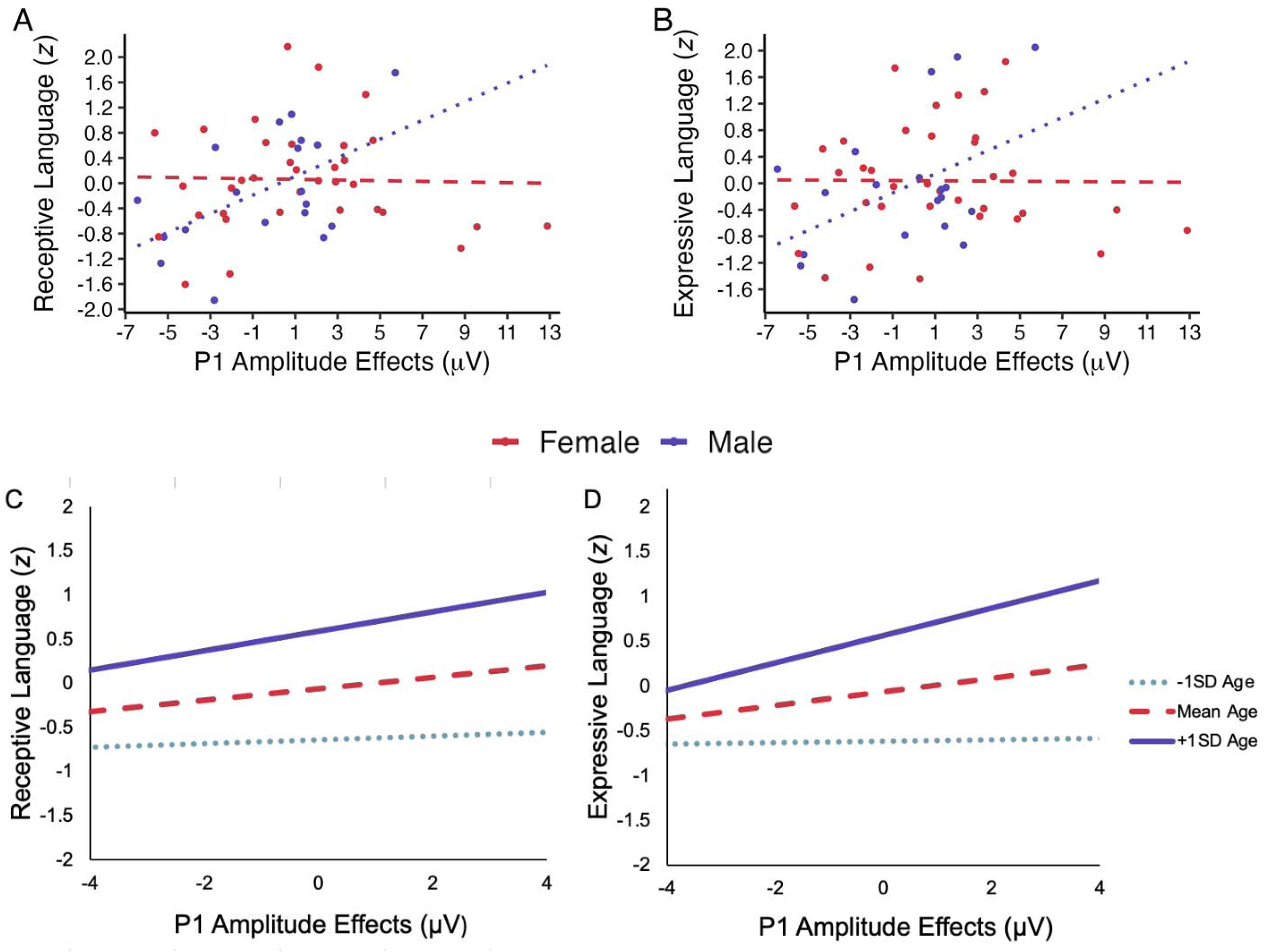
Associations Between P1 Amplitude Effects and Receptive and Expressive Language According to Chronological Age and Biological Sex. *Note*. Plots depicting associations for P1 amplitude effects and receptive and expressive language aggregate *z* scores. Panels A and B depict associations varying by biological sex: red = females; blue = males. Greater P1 amplitude effects were associated with greater receptive and expressive language in males, with negligible associations in females. Panels C and D depict associations varying by chronological age: dotted gray line = -1 *SD* age, dashed red line = mean age, solid blue line = +1 *SD* age. Associations between P1 amplitude effects and language were significant for infants over 14 months of age.

Zero-order correlations between N2 amplitude effects and receptive and expressive language were not significant, *r* = .170, *p* = .219 and *r* = .107, *p* = .441, respectively. These non-significant relations between N2 amplitude effects and language were small in magnitude. Associations between N2 amplitude effects and receptive and expressive language were not significantly moderated by group, chronological age, or biological sex (*p* values for interaction terms in models testing moderated effects > .1).

## Discussion

This study is the first to our knowledge to identify differential ERP amplitudes in response to audiovisual versus auditory-only speech in infants at elevated and general-population-level likelihood for autism. Additionally, these ERP amplitude effects were linked to looking to audiovisual speech as measured by eye tracking, as well as to concurrent receptive and expressive language ability, for at least some participants. Overall, these findings indicate that looking to and processing of audiovisual speech early in life may be relevant for language development and, thus, have important implications for autistic people’s outcomes.

### ERP Amplitudes Differ in Response to Audiovisual versus Auditory-Only Speech

N2 amplitudes were significantly more negative in response to audiovisual versus auditory-only speech, on average, across groups. This result extends prior work identifying P2 amplitude differences in response to audiovisual versus auditory-only speech in school-aged autistic and non-autistic children (Dunham-Carr et al., 2023) and suggests that differential processing of multisensory speech as indexed by ERPs is present earlier in life than had previously been demonstrated, within 12–18-months. Both P2 in older children and adults and N2 in infants in response to speech have been thought to reflect processing of phonemes (Baart et al., 2014; Kaganovich & Schumaker, 2014). Therefore, prior findings in school-aged children and these findings in infants, taken together, suggest that P2 and N2 amplitude effects reflect audiovisual phonetic processing.

There were no mean group differences in ERP amplitudes in response to audiovisual versus auditory-only speech in infants at high and low likelihood for autism diagnosis. This finding is not surprising, perhaps, given that we failed to find between-group differences in our prior study investigating ERP responses to audiovisual versus auditory-only speech in school-aged autistic and non-autistic children (Dunham-Carr et al., 2023). Notably, across both school age children and infants, we observed considerable individual variability in ERP amplitudes within and across diagnostic groups that mapped onto concurrent language-related metrics. These findings collectively suggest that individual differences in ERP responses to audiovisual speech may be more informative than mean differences between groups.

P1 amplitudes did not significantly differ across conditions. Psychometric limitations may explain the lack of condition-related differences in P1 amplitudes. In school-aged autistic children, amplitudes for N1 (the negative ERP deflection commonly seen in approximately the same time window as the infant P1 in older children and adults) were found to be unstable, possibly due to differences in the maturation of the N1 component in the age range sampled (Dunham et al., 2020). It is possible that differences in waveform composition across early childhood may similarly contribute variability in P1 amplitudes, impacting the ability to detect effects of interest (e.g., Sharma et al., 1997; Wunderlich & Cone-Wesson, 2006). Further psychometric work investigating the stability and validity of ERP metrics in response to auditory-only and audiovisual speech, as well as ERP metrics in infants, is needed.

### Looking to Audiovisual Speech is Associated with ERP Amplitude Effects

Looking to audiovisual speech was associated with ERP amplitude effects but varied significantly according to sibling group. The significant group moderations suggest that the direction and magnitude of links between looking to and processing of audiovisual speech differ for Sibs-Autism versus Sibs-NA. For example, increased looking to the mouth covaried with greater P1 and lesser N2 amplitude effects in Sibs-Autism, but the opposite trends were observed in Sibs-NA.

Links between looking to the mouth of a speaker and ERP amplitudes to audiovisual speech have also been identified in infants in prior studies. For example, Kushnerenko et al. (2013) found that the amplitude of a mismatch response to incongruent audiovisual speech stimuli was negatively associated with 6-9-month-old infants’ looking time to the mouth, such that a shift to increased looking at the mouth of the speaker corresponded with less of an audiovisual mismatch response. It is possible that the differential associations in looking to audiovisual speech and ERP amplitude effects that we have observed in Sibs-Autism and Sibs-NA reflect different developmental trajectories in looking to and processing of audiovisual speech. These findings provide increased empirical support for the notion that looking to audiovisual speech is linked with audiovisual speech processing but underscore the need for future investigations to consider how and why these associations may vary by familial predisposition for autism.

### ERP Amplitude Effects Are Associated with Emerging Language Ability

Amplitude effects, in particular at P1, were also associated with emerging language ability in our sample, when accounting for participant characteristics. These findings expand upon prior work identifying links between P2 amplitude differences and vocabulary in school-age children, for whom P2 amplitude suppression for audiovisual relative to auditory-only speech was associated with expressive vocabulary scores through receptive vocabulary scores (Dunham-Carr et al., 2023). Here, greater P1 amplitude effects (i.e., more P1 amplitude suppression in response to audiovisual speech compared to auditory-only speech) corresponded with greater receptive and expressive language ability for some participants, specifically (a) males and (b) infants older than 14 months of age. Overall, our findings provide some support for our hypothesis that ERP amplitude effects may explain heterogeneity in concurrent language ability but emphasize again that such effects may only explain variability in language for a subset of participants.

### Considering Participant Characteristics in Analyses

It is important to note that we would not have detected associations between looking, audiovisual speech processing, and early language ability if we had not considered participant characteristics with the potential to influence the relations of interest, all of which varied according to at least one putative moderator. These findings add to a growing literature on correlations between behaviors of interest to autism researchers in infants that differ according to participant-level factors. Our findings are particularly consistent with prior investigations of other aspects of early sensory processing and behavior in Sibs-Autism and Sibs-NA that found associations moderated by sibling group (e.g., Damiano-Goodwin et al., 2018; Feldman et al., 2024; Wagner et al., 2018) and chronological age (e.g., Feldman et al., 2021; Gunderson et al., 2024).

Sex differences in patterns of looking to audiovisual speech stimuli and language have also been documented in autistic children and in infants at elevated likelihood for an autism diagnosis (e.g., Chawarska et al., 2016; Harrop et al., 2020; see Lozano et al., 2025 for a review). Notably, sex differences in prevalence and features have been identified across neurodevelopmental conditions, and biological sex influences neural and language development in a myriad of ways, including sex-differential patterns across brain anatomy and function, sex hormone impacts on neurotransmitter systems and synapse formation, and differences in communication related behaviors during language acquisition (Bölte et al., 2023). Further research investigating sex-related factors of neurodevelopment is critical for elucidating mechanisms underpinning sex differences in neural and language development. Overall, it is important for researchers to not only consider, but also systematically assess these empirically motivated characteristics *a priori* to permit their consideration as moderators in analyses.

### Strengths and Limitations

This study provides further empirical support for the cascading effects framework by demonstrating that looking to audiovisual speech as measured by eye tracking, as well as differential audiovisual speech processing as indexed by ERP amplitude effects, explain heterogeneity in emerging language ability in infants at increased and general population-level likelihood for autism. We recruited a sample of Sibs-Autism and Sibs-NA groups that did not significantly differ on age and biological sex at study entry while also yielding a study sample with a high degree of heterogeneity in language ability across groups, facilitating our detection of associations of interest.

However, this study is not without limitations. First, our sample was predominantly White and non-Hispanic, limiting the generalizability of our findings to infants from other racial and ethnic backgrounds. Additional work is needed to explore links for looking, audiovisual speech processing, and language ability in more representative samples, especially in children from demographic groups that are historically underrepresented in scientific research and that may experience different language learning environments that may influence effects of interest. Given the exploratory nature of this investigation and the relatively small sample size, it is also important to note that several regression analyses were carried out without correcting for multiple comparisons, which increases the risk of family-wise error. Finally, we only evaluated associations between looking to audiovisual speech, audiovisual speech processing, and receptive and expressive language through a concurrent correlational design. As a result, we cannot presently draw any conclusions regarding the directionality or causality of the observed associations. Future larger-scale studies incorporating longitudinal and/or experimental designs with a broader range of audiovisual speech stimuli are needed to better understand precisely how early looking to and processing of audiovisual speech influences language development and outcomes across Sibs-Autism and Sibs-NA.

### Conclusions

This study extends our team’s past findings of ERP amplitude effects in response to audiovisual versus auditory only speech to 12-18-month-old infants with and without autistic older siblings. The results demonstrate that differential ERP amplitudes for audiovisual relative to audiovisual speech are present and associated with looking to audiovisual speech and emerging language at 12-18 months of age but underscore the fact that participant characteristics must be considered. These findings support further investigation of the role of audiovisual speech processing in early language development on a larger scale and suggest that early interventions targeting looking to and processing of audiovisual speech may be useful for optimizing the language and long-term outcomes of infants at increased and general population level likelihood for autism and developmental language disorder.

## Acknowledgments

The authors would like to thank the Adventure Science Center in Nashville, TN for their Living Laboratory partnership, as well as the participants and their families.

## Author Contributions

Kacie Dunham-Carr conceptualized the research project, conducted data collection, processing, and analyses, and drafted the manuscript. Bahar Keçeli-Kaysılı, Jennifer Markfeld, Grace Pulliam, S. Madison Clark, and Jacob I. Feldman conducted data collection and assisted with data processing. Pooja Santapuram developed eye tracking data extraction methods and assisted with data collection and processing. Kelsea McClurkin, Drina Agojci, and Ava Schwartz assisted with data collection and processing. David J. Lewkowicz developed eye tracking stimuli and protocol. Tiffany G. Woynaroski conceptualized the research project, obtained research funding, and oversaw data collection, processing, and analyses. All authors contributed to the editing and approval of the drafted manuscript.

## Statements and Declarations

### Ethical Considerations

Procedures were approved by the Institutional Review Board at Vanderbilt University Medical Center.

### Consent to Participate

Parents of participants provided written informed consent prior to participation in the study.

### Data Availability Statement

Data collection for the larger project of which this study is a part of is still ongoing; thus, data are not publicly available at this time. Data may be available upon request to the authors at study conclusion.

## Footnotes

1 In this manuscript, we have elected to use identity-first language to reflect the current wishes of the autistic community and recent recommendations on terminology for autism researchers (Bottema-Beutel et al., 2021).

2 The older sibling of one participant declined to participate in diagnostic confirmation for the study. Group level analyses were robust to excluding this participant, and this participant was not identified as an outlier in regression analyses.

3 Social Communication Questionnaires for three older siblings in the Sibs-NA group were not completed due to protocol disruptions during the COVID-19 pandemic. Group level analyses were robust to excluding these participants, and they were not identified as outliers in regression analyses.

