## Supplement for "Looking to and Processing of Audiovisual Speech and Associations with Language in Infant Siblings of Autistic and Non-autistic Children"

**SUPPLEMENTAL MATERIALS**

**Supplemental Figure 1**

face

eyes

mouth

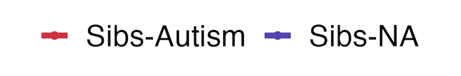
*Associations Between Looking to Eyes and ERP Amplitude Effects*

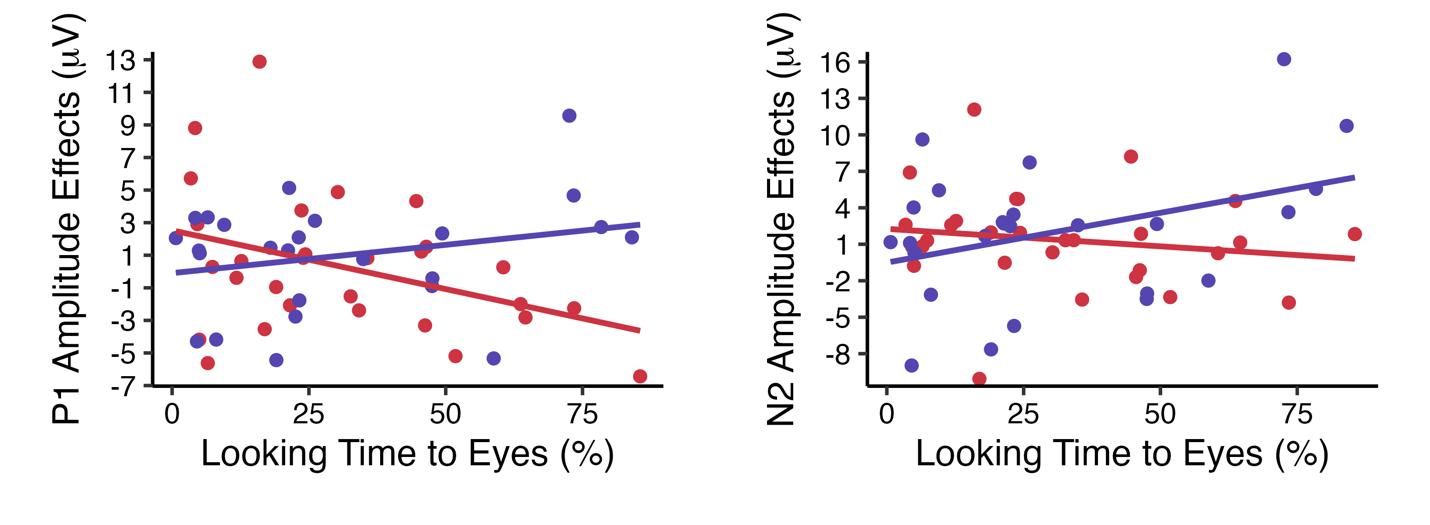

*Note.* Scatterplots depicting associations between looking to the eyes of the speaker and P1 and N2 amplitude effects. Associations are moderated by group. Red = Sibs-Autism group; blue = Sibs-NA group.

**Supplemental Figure 2**

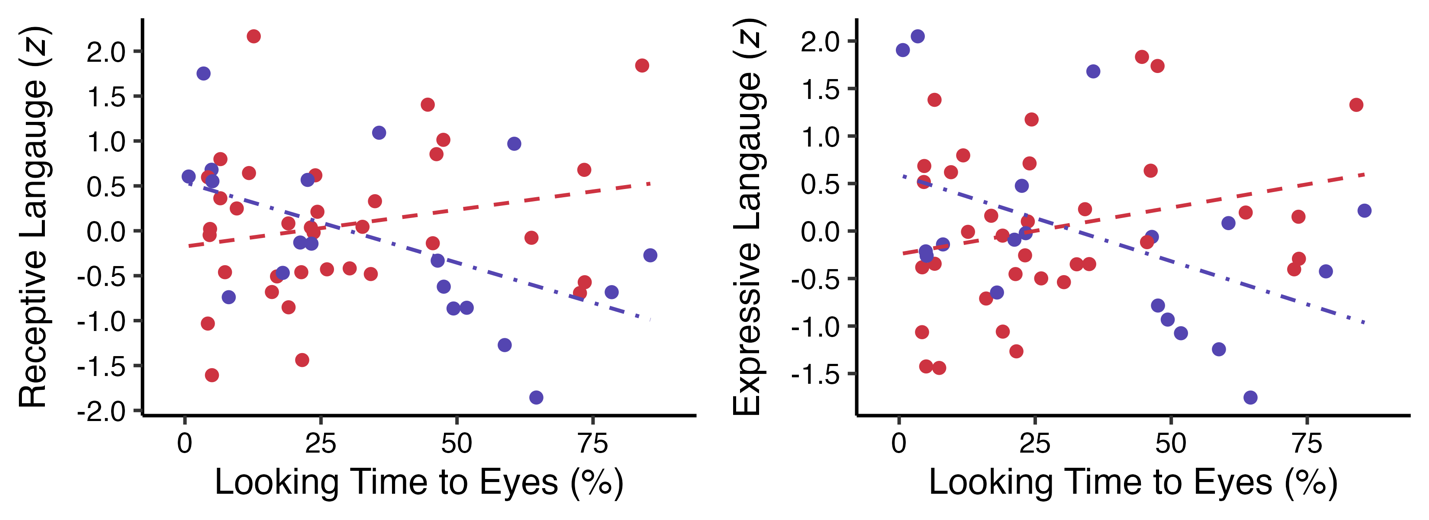
*Associations Between Looking to Eyes and Receptive and Expressive Language*

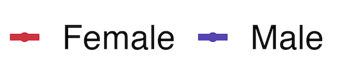

*Note.* Scatterplots depicting associations between looking to the eyes of the speaker and receptive and expressive language. Associations are moderated by biological sex. Red = females, blue = males.

**Supplemental Table 1**

*Zero-Order Correlations Between Component Variables Purported to Tap Receptive and Expressive Language*

| Receptive Language Variables | 1 | 2 | 3 |  |  |  |
| --- | --- | --- | --- | --- | --- | --- |
| 1. Mullen Receptive Age Equivalency Score | — |  |  |  |  |  |
| 2. VABS Receptive Age Equivalency Score | .713*** | — |  |  |  |  |
| 3. MCDI Understands Raw Score | .429** | .624*** | — |  |  |  |
| Expressive Language Variables |  |  |  | 4 | 5 | 6 |
| 4. Mullen Expressive Age Equivalency Score |  |  |  | — |  |  |
| 5. VABS Expressive Age Equivalency Score |  |  |  | .686*** | — |  |
| 6. MCDI Says Raw Score |  |  |  | .726*** | .756*** | — |

*Note.* Intercorrelations between component variables purported to tap receptive and expressive language, respectively. Prior to evaluating intercorrelations, the Mullen Receptive Language Age Equivalency score and MCDI Understands Raw score were square root transformed, and the MCDI Says Raw Score was log transformed to correct for slight positive skew. VABS = Vineland Adaptive Behavior Scales, second edition (Sparrow et al., 2005); MCDI = MacArthur-Bates Communicative Development Inventories (Fenson et al., 2007); Mullen = Mullen Scales of Early Learning (Mullen, 1995).

**p* <.05, ***p* < .01, ****p* < .001.

**Supplemental Table 2**

*Zero-Order Correlations Across All Variables of Interest to Analyses*

| Variable | 1 | 2 | 3 | 4 | 5 | 6 |
| --- | --- | --- | --- | --- | --- | --- |
| 1. Expressive Language | — |  |  |  |  |  |
| 2. Receptive Language | .763*** | — |  |  |  |  |
| 3. P1 amplitude effects | .147 | .158 | — |  |  |  |
| 4. N2 amplitude effects | .107 | .170 | .707*** | — |  |  |
| 5. Looking to mouth | .094 | .112 | .035 | -.215 | — |  |
| 6. Looking to eyes | -.092 | -061 | -.100 | .145 | -.828*** | — |

*Note.* Receptive and expressive language are indexed here by aggregate variables formed by averaging the component variables summarized above in Supplemental Table 4.1, following confirmation of sufficient intercorrelation and z-score transformation.

**p* <.05, ***p* < .01, ****p* < .001.

**Supplemental Table 3**

*Results for Regression Analyses Testing Relations Between Looking to Audiovisual Speech and P1 Amplitude Effects*

|  |  | Unstandardized Coefficients | | Standardized Coefficients | |  |  |
| --- | --- | --- | --- | --- | --- | --- | --- |
| Model | Regressor | B | SE | Beta | *t* | *p* | *f^2^* |
| 1 | Constant  Looking to Eyes | 1.09  -.016 | .882  .022 | —  -.100 | 1.23  -.725 | .223  .472 | —  .010 |
| 2 | Constant  Looking to Mouth | .343  .005 | 1.08  .018 | —  .035 | .316  .254 | .753  .801 | —  .001 |
| 3 | Constant  Looking to Eyes  Group  Looking to Eyes x Group | 5.15  -.179  -2.63  .107 | 2.70  .069  1.69  .043 | —  -1.10  -.336  1.13 | 1.91  -2.59  -1.55  2.48 | .062†  .013*  .127  .017* | —  .134  .048  .123 |
| 4 | Constant  Looking to Mouth  Group  Looking to Mouth x Group | -7.47  .132  5.16  -.084 | 3.31  .055  2.08  .034 | —  1.03  .660  -1.20 | -2.26  2.42  2.48  -2.46 | .028*  .019*  .017*  .017* | —  .117  .123  .121 |

*Note.* Coefficients, *p* values, and *f*^2^ for regression analyses. According to Cohen (1988), *f^2^* values of .02, .15, and .35 correspond to small, medium, and large effect sizes.

†*p* value for effect <.1, **p* value for effect < .05.

**Supplemental Table 4**

*Results for Regression Analyses Testing Relations Between Looking to Audiovisual Speech and N2 Amplitude Effects*

|  |  | Unstandardized Coefficients | | | | Standardized Coefficients | |  | | |
| --- | --- | --- | --- | --- | --- | --- | --- | --- | --- | --- |
| Model | Regressor | B | | SE | | Beta | *t* | | *p* | *f*^2^ |
| 1 | Constant  Looking to Eyes | .748  .029 | 1.08  .028 | | .145 | | .691  1.06 | | .492  .296 | —  .021 |
| 2 | Constant  Looking to Mouth | 3.46  -.034 | 1.31  .021 | | -.215 | | 2.64  -1.59 | | .011*  .118 | —  .049 |
| 3 | Constant  Looking to Eyes  Group  Looking to Eyes x Group | 5.04  -.139  -2.77  .111 | 3.37  .087  2.12  .054 | | -.694  -.288  .947 | | 1.49  -1.61  -1.32  2.05 | | .141  .113  .196  .045* | —  .052  .034  .084 |
| 4 | Constant  Looking to Mouth  Group  Looking to Mouth x Group | -4.23  .093  5.08  -.083 | 4.07  .067  2.57  .042 | | —  .589  .527  -.967 | | -1.04  1.38  1.98  -1.99 | | .303  .173  .053†  .053† | —  .038  .078  .079 |

*Note.* Coefficients, *p* values, and *f*^2^ for regression analyses. According to Cohen (1988), *f^2^* values of .02, .15, and .35 correspond to small, medium, and large effect sizes.

†*p* value for effect <.1, **p* value for effect < .05.

**Supplemental Table 5**

*Results of Regression Analyses Testing Relations Between Looking to Audiovisual Speech and Receptive and Expressive Language*

| Associations with Receptive Language | | | | | | | |
| --- | --- | --- | --- | --- | --- | --- | --- |
| Model |  | Unstandardized Coefficients | | Standardized Coefficients | |  |  |
|  |  | B | SE | Beta | *t* | *p* | *f*^2^ |
| 1 | Constant  Looking to Eyes | .101  -.003 | .191  .005 | -.092 | .529  -.668 | .599  .507 | —  .009 |
| 2 | Constant  Looking to Mouth | -.139  .003 | .234  .004 | .094 | -.592  .683 | .556  .498 | —  .009 |
| 3 | Constant  Looking to Eyes  Biological Sex  Looking to Eyes x Biological Sex | 1.25  -.044  -.713  .026 | .675  .016  .389  .009 | -1.25  -.403  1.25 | 1.85  -2.81  -1.84  2.76 | .070†  .007**  .072†  .008** | —  .016  .067  .152 |
| 4 | Constant  Looking to Mouth  Biological Sex  Looking to Mouth x Biological Sex | -1.23  .020  .710  -.011 | .758  .013  .468  .008 | .730  .402  -.793 | -1.63  1.54  1.52  -1.43 | .110  .131  .136  .160 | —  .047  .046  .041 |
| Associations with Expressive Language | | | | | | | |
| 5 | Constant  Looking to Eyes | .071  -.002 | .203  .005 | -.061 | .347  -.438 | .730  .663 | —  .004 |
| 6 | Constant  Looking to Mouth | -.175  .003 | .248  .004 | .112 | -.705  .813 | .484  .420 | —  .013 |
| 7 | Constant  Looking to Eyes  Biological Sex  Looking to Eyes x Biological Sex | 1.42  -.046  -.833  .028 | .718  .017  .413  .010 | -1.23  -.445  1.26 | 1.98  -2.78  -2.02  2.78 | .053†  .008**  .049*  .008** | —  .154  .081  .155 |
| 8 | Constant  Looking to Mouth  Biological Sex  Looking to Mouth x Biological Sex | -1.73  .033  1.01  -.019 | .780  .014  .482  .008 | 1.11  .537  -1.23 | -2.22  2.41  2.09  -2.29 | .031*  .020*  .042*  .027* | —  .117  .087  .104 |

*Note.* Coefficients, *p* values, and *f*^2^ values. According to Cohen (1988), *f^2^* values of .02, .15, and .35 correspond to small, medium, and large effect sizes.

†*p* value for effect <.1, **p* value for effect < .05, ***p* value for effect < .01.

**Supplemental Table 6**

*Results for Regression Analyses Testing Relations between P1 Amplitude Effects and Receptive and Expressive Language According to Chronological Age and Sex*

| Associations with Receptive Language | | | | | | | |
| --- | --- | --- | --- | --- | --- | --- | --- |
| Model |  | Unstandardized Coefficients | | Standardized Coefficients | |  |  |
|  |  | B | SE | Beta | *t* | *p* | *f*^2^ |
| 1 | Constant  P1 Amplitude Effects | -.020  .034 | .117  .030 | .158 | -.171  1.57 | .865  .252 | —  .026 |
| 2 | Constant  P1 Amplitude Effects  Chronological Age  P1 Amplitude Effects x Chronological Age | -3.61  -.204  .234  .0177 | .592  .139  .038  .010 | -.940  .631  1.15 | -6.10  -1.50  6.12  1.80 | <.001***  .148  <.001***  .077 | —  .043  .749  .065 |
| 3 | Constant  P1 Amplitude Effects  Biological Sex  P1 Amplitude Effects x Biological Sex | -.148  .302  .106  -.153 | .408  .124  .239  .069 | 1.39  .060  -1.28 | -.362  2.42  .442  -2.23 | .719  .019*  .661  .030* | —  .117  .004  .100 |
| Associations with Expressive Language | | | | | | | |
| 4 | Constant  P1 Amplitude Effects | -.020  .034 | .124  .032 | .147 | -.159  1.07 | .875  .288 | —  .022 |
| 5 | Constant  P1 Amplitude Effects  Chronological Age  P1 Amplitude Effects x Chronological Age | -3.44  -.338  .223  .027 | .650  .152  .042  .011 | -1.47  .567  1.67 | -5.29  -2.22  5.31  2.53 | <.001***  .031*  <.001***  .015* | —  .099  .564  .128 |
| 6 | Constant  P1 Amplitude Effects  Biological Sex  P1 Amplitude Effects x Biological Sex | -.045  .286  .041  -.144 | .439  .134  .257  .074 | 1.24  .022  -1.13 | -.102  2.14  .160  -1.95 | .919  .038*  .874  .057† | —  .091  .001  .076 |

*Note.* Coefficients, *p* values, and *f*^2^ values for regression analyses. According to Cohen (1988), *f^2^* values of .02, .15, and .35 correspond to small, medium, and large effect sizes.

†*p* value for effect <.1, **p* value for effect < .05, ***p* value for effect < .01, ****p* value for effect < .001.
